# The Cerebellum Plays More Than One Role in the Dysregulation of Appetite: Review of Structural Evidence from Typical and Eating Disorder Populations

**DOI:** 10.1101/2022.04.14.22273867

**Authors:** Michelle Sader, Gordon D. Waiter, Justin H. G. Williams

## Abstract

**Objective:** Dysregulated appetite control is characteristic of anorexia nervosa (AN), bulimia nervosa (BN) and obesity (OB). Studies using a broad range of methods suggest the cerebellum plays an important role in aspects of weight and appetite control, and is implicated in both AN and OB by reports of aberrant grey matter volume (GMV) compared to non-clinical populations. As functions of the cerebellum are anatomically segregated, specific localization of aberrant anatomy may indicate the mechanisms of its relationship with weight and appetite in different states. We sought to determine if there were consistencies in regions of cerebellar GMV changes in AN/BN and OB, as well as across normative variation.

**Method:** Systematic review and meta-analysis using GingerALE.

**Results:** Twenty-six publications were identified as either case-control studies (nOB=277; nAN/BN=510) or regressed weight from normative (NOR) data against brain volume (total n=3,830). AN/BN and OB analyses both showed consistently decreased GMV within Crus I and Lobule VI, but volume reduction was bilateral for AN/BN and unilateral for OB. Analysis of the normative dataset identified a cluster in right posterior lobe which overlapped with AN/BN cerebellar reduction. Sensitivity analyses indicated robust repeatability for NOR and AN/BN cohorts, but found OB-specific heterogeneity.

**Discussion:** Findings suggest that more than one area of the cerebellum is involved in control of eating behaviour and is differentially affected in normal variation and pathological conditions. Specifically, we hypothesise an association with sensorimotor and emotional learning via Lobule VI in AN/BN, and executive function via Crus I in OB.

## 1.0 Introduction

### 1.1 Importance of Studying Appetite Control Mechanisms

Appetite control has a complex and multifaceted nature, and problems with eating behaviour arise from a variety of genetic, cognitive, emotional and physiological factors. Irregular appetite patterns result in abnormal body weight for height (indexed by the Body Mass Index – BMI) as well as metabolic and mental health^1–3^.

Obesity (OB), characterised by a BMI of >30, is a major public health concern pertaining to appetite dysregulation that is on the increase and constitutes as a major risk factor for conditions such as hypertension, diabetes, cardiovascular diseases and cancer^4–6^. In the past 35 years, worldwide OB prevalence rates have nearly doubled. Currently, 15% and 11% of women and men respectively were classified with OB, and approximately 42 million children <5 years were classified as having excess weight (EW)^7^. It is a priority for research to identify mechanisms that control appetite which may serve as a treatment targets.

At the opposite end of the BMI scale are individuals with anorexia nervosa (AN) who refrain from eating and harbour pathological fears of weight gain and food consumption^8^. AN is a complex multidimensional eating disorder characterised by pathologically decreased weight-for-age/height, with lifetime prevalence as high as 4%^9^. Recently, publications found an increase in reported AN cases over time, although increased incidence may correlate with increased specificity of reporting protocols^10,11^. While AN is relatively uncommon compared to other psychiatric disorders, mortality rates are greater, reporting between 2-6%^12,13^. While risk of AN development is far lower than risk OB development, AN serves as a vital example of consequences related to appetite dysregulation.

Whilst AN and OB may not technically be eating disorders at opposite ends of a singular spectrum, there are prominent neuroanatomical^14–26^, metabolic and genetic^27–31^ factors linking both disorders. An intriguing possibility is whether common neurobiological or neuroanatomical mechanisms could be implicated in both OB and AN. Both conditions present dysregulation of BMI/appetite, with individuals prone to engaging in behaviours that exacerbate pathological weight increase/decrease. As OB and AN are so disparately associated with body weight, such mechanisms likely function in condition-dependent manners, or that similar, rather than identical mechanisms underlie both conditions.

### 1.2 A Cerebellar Role in Weight & Appetite Regulation

Cerebellar roles (displayed in *Figure S1*^32^) in body-weight or appetite control^33–35^ receive surprisingly little attention in appetite-related research. Traditionally, this brain region was thought to solely serve motor coordination and somatic functions, but further investigation reveals that the cerebellum plays varieties of diverse roles. Researchers have reported the cerebellum demonstrates organisational similarity to that of the cerebral cortex^36^ with anatomically segregated functions. Cerebellar contributions to intrinsic connectivity networks have shown that particular regions of the cerebellum are distinctly involved in different cognitive functions^37–39^ and is implicated in five intrinsic connectivity networks, including the executive control network ([ECN] via Lobule VIIB; Crus I/II), default-mode network (via Lobule IX), salience network (via Lobule VI), and sensorimotor network (via Lobule VI)^37^. Contrasting traditional conceptualization, motor tasks are largely consigned to Lobule VIIIa/b and represent limited cerebellar functionality^39^.

Current views of the cerebellum implicate it in homeostatic regulation^40,41^, executive/cognitive functionality^37,39,42–47^ (including habit formation^45^, conditioning behaviours^37,45,48–51^, procedural knowledge storage^37,48,49,52^, working memory function^39,51^ and cravings^53^), and emotional regulation^37,39,46,49,50,54–58^. Importantly, evidence suggests the cerebellum may participate in aspects of food intake and appetite control through multiple mechanisms, including physiological (i.e., feeding circuit connectivity, response/influence on gut hormones/neurotransmitters), cognitive (i.e., food palatability, feeding-related memories) and emotional (i.e., food-related cravings) means^59,60^.

On a physiological scale, the cerebellum interacts via extensive signalling networks with the hypothalamus and insula, which both contain networks specific to food intake^42,60^ via neural and hormonal mechanisms^14,42,61^. Enteric nervous system gut hormones, such as leptin and ghrelin, interactively modulate regions of the brain associated with food intake control including the cerebellum, hypothalamus and brainstem^62,63^. In response to ghrelin, cerebellar activation decreases and likely works to stimulate appetite via ghrelin-induced suppression of satiety hormones, such as cholecystokinin, within vagal afferent neurons^64–66^. Recently, Choe et al. (2021)^67^ investigated synchrony between gastric rhythm and brain activity, identifying the strongest synchrony within the cerebellum^67^, suggesting a more direct relationship between gastric mechanisms and cerebellar functionality.

The cerebellum is also implicated in genetic aspects associated with bodyweight disorders. Beyond neuroanatomical/biological aspects of appetite dysregulation, disorders on the extremes of the standard BMI-measure (i.e., AN/OB) exhibit shared genetic and metabolic correlations^27–30^ to the extent with which they have been termed metabolic “mirror images” of one another^31^. Cerebellar tissues and pathways have recently been implicated in aspects of genetic risk for both conditions^27,68^. Evidence from AN- and cerebellum-related studies suggest that deficits in cerebellar mRNA expression occur in foetal and early-life AN pathogenesis^68^, and altered cerebellar volume may explain body image disturbances in AN^27,69–71^. In OB, a multitude of genes have been associated with increased risk, predominantly the fat mass (FTO) and melanocortin-4 receptor genes. FTO is expressed in regions such as the hypothalamus, hippocampus and cerebellum^72–75^, and research suggests it is highly associated with OB outcome^28,29^.

The cerebellum is also repeatedly reported to participate in neurobiological modulation of dopaminergic and serotonergic signalling^76,77^, which becomes dysregulated in conditions of abnormal weight/BMI^78–83^. Dopaminergic-related mechanisms driving both OB and AN have also been associated with addiction-resembling behaviour related to eating behaviour^84–86^. Recently, Low *et al.* (2021)^87^ used a reverse-translational approach to identify a cerebellar-based satiation network. In humans, food cues activate cerebellar output neurons to promote satiation through reductions in phasic dopaminergic responses to food. As such, the cerebellum is likely implicated in altered neurobiological mechanisms associated with dysregulated appetite or weight, such as those with AN/OB.

### 1.3 Cerebellar Volume and Dysregulation of Appetite

Structural associations between the cerebellum and conditions of dysregulated BMI and appetite have been consistently documented. This includes evidence from loss-of-function studies^88^, as well as animal studies where lesions or removal of cerebellar hemispheres leads to reduced appetite, pathological weight loss and increased mortality rate^89^. In humans, Oya *et al*. (2014)^90^ reports a high proportion of cerebellar tumour detection with associated AN. Similarly, cerebellar degeneration associated with ataxia correlates with increased likelihood of being underweight or experiencing abnormal appetite^93–9691–94^. Due to appetite disturbances emerging post-cerebellar structural abnormalities, these studies indicate that cerebellar deficits could play a causative role in the loss of appetite.

Cerebellar volume has also been directly associated with conditions of under- and over-eating. Excess weight (BMI>25.5) or OB (BMI>30.0) has been associated with both increased^16^ and decreased concentrations of grey matter volume (GMV) in the left^16^, bilateral^15^ and bilateral posterior^19^ cerebellum. Prefronto-cerebellar circuits, implicated in cognition, emotion, executive function and error detection, exhibit volume reduction in those with OB^14,16^. Recovery from OB is also associated with positive changes in cerebellar volume, and is thought to be important in treatment as well as conditioning of eating behaviour^20^. Evidence suggests affected cerebellar regions in excess weight play roles in the storage of procedurally learned knowledge^16^, which may influence food consumption and emotional eating behaviours via the alteration or dysfunction of food-related learning strategies and tasks, particularly concerning knowledge on healthy dietary practices. Alternatively, the cerebellum may play a role in appetite during states of OB/excess weight via functional alterations in hypothalamic networks^17,18^. Differences in cerebellar volume have been noted in both rat and human models of AN^84,95,96^. Within humans, cerebellar atrophy and cellular loss is associated with AN disease duration, poor treatment success^26^, persists post weight-recovery, and is suggested to play a role in maintaining a low body weight^21,22^.

### 1.4 A Cerebellar Role in Volitional Appetite Control?

Extant literature also suggests that the cerebellum may participate in more conscious domains of appetite, such as emotional and cognitive aspects driving appetitive behaviours. The cerebellum may play a substantive role in cognitive aspects of appetite control via an individuals’ subjective feeling of craving or outcome expectation built off previous experience and memory. This region has been a focal point of research into the experience of craving, with studies using drug- and food-related stimuli to assess cerebellar effects^53^. In those with OB as well as binge-eaters, greater connectivity between the cerebellum and anterior cingulate cortex was observed during food-related cues^53,97,98^. Studies using cocaine-/tobacco-related cues demonstrate similarly increased activation and effects overlapped with those seen in food-related cue studies^99,100^. A meta-analysis^101^ examining brain activation across food/drug/sex/gambling cues identified cerebellar activation to all cue types, including food-related cues.

The cerebellum is also associated with traits of impulsivity and loss-of-control (LOC) eating. Recent investigations unveiled cerebellar roles in the portion size effect (PSE)^102–104^, the phenomenon where more is eaten when large quantities of food are available. English *et al.* (2019)^105^ measured traits of impulsivity including LOC eating in association with regional brain activation, reporting increased activation of the left cerebellum when adolescent LOC participants viewed pictures of food items with high vs. low energy density. This mechanism has been proposed to play a role in the PSE, appetite dysregulation and the development of obesity^105^. Interestingly, findings regarding a reduction of cerebellar volume in those with OB contrast reports of increased activity during food-/drug-related tasks and in LOC eating, which may suggest multiple cerebellar structures differentially contribute to OB or excess consumption. In those with AN relative to controls, cerebellar and prefrontal hyperactivation was reported during response inhibition in a physical activity paradigm and suggested to contribute to altered executive function and inhibitory control^106^.

Altogether, the cerebellum may function as a pivotal point of behavioural tuning in relation to the cognitive aspects of appetite regulation. Schmahmann *et al*. (2019)^107^ described associations between the posterior lobe and cognition and proposed “The Dysmetria of Thought” theory. This theory suggests that the cerebellum modulates cognition in a similar way to which it smooths and fine-tunes coordination of motor behaviour by receiving multimodal inputs and establishing procedurally optimized behavioural modulation, similar to that observed in motor control and likely to occur in connection with the prefrontal cortex. The cerebellum may perform similarly in regards to the input and integration of appetite-related stimuli, such as individualised food-related memories and expectations. Concerning restrictive eating and low BMI reported in AN, it is possible that cerebellar regions attending to salience of food stimuli, such as Lobule VI, may be associated with changes in bodyweight/appetite via formations of negative associations with food. Alternatively, cognitive/executive abnormalities reported in those with OB may suggest associations with cerebellar regions prominent to the ECN, such as Crus I, where reward value or experiential engagement with food stimuli may be altered. As both conditions, regardless of disparate body-weight states, also report with altered emotional/cognitive traits, it is also possible that both salience- and executive-associated cerebellar circuits are implicated in AN and OB but in condition-dependent manners.

### 1.5 Aims of Study

Evidence provided by wide-ranging cerebellar research demonstrates that the cerebellum is important in modulating aspects of appetite control and implicated in both OB and AN. However, whether identical or differing areas of the cerebellum are associated with AN and OB are unclear. This systematic review aimed to examine cerebellar anatomy reported in case-control AN and OB literature, as well as across the weight range via normative/non-clinical populations to investigate whether cerebellar structure differs across body-weight states and disorders. We generated two alternative hypotheses: We first hypothesised that a singular area of the cerebellum, such as Crus I (implicated in executive control circuits^20^), would be associated with abnormal BMI conditions, and that respective regions affected in OB and AN would be similar. Our alternative, competing hypothesis, was that separate cerebellar regions would be altered across conditions and shown in distinct regions. For instance, Crus I volume could be primarily affected in OB, while Lobule VI volume (implicated in salience/sensorimotor circuits) could be primarily affected in AN.

## 2.0 Methodology

### 2.1 Selection of Literature

Literature was searched on June 29, 2022 using SCOPUS, PubMed Central (PMC), and Web of Science (WoS) using identical search criteria. SCOPUS identified 2,638 publications involving cerebellar volume during OB [search criteria: [*(cerebell* AND obesity AND MRI)*] and 572 publications regarding AN [*(cerebell* AND anorexia AND MRI)*]. PMC identified 4,950 publications relating to OB, as well as 461 papers regarding AN. Lastly, 2,588 and 233 papers were identified through WoS regarding cerebellar characteristics in those with OB and AN respectively. Inclusion criteria involved presence of key phrases utilised in search, including “grey literature”, or findings produced outside of traditional publishing. Exclusion criteria were as follows: 1.) Publications older than 10 years, as CB imaging methods have significantly improved since 2010^108–110^; 2.) Animal studies; 3.) Publications not reporting case-control studies 4.) Inclusion of clinical groups/correlations unrelated to the study scope; 5.) Publications not utilising voxel-based morphometry (i.e., DTI/fMRI studies); 6.) Publications with results not reported in Talairach/MNI coordinates. Individual studies fitting criteria were additionally collected from meta-analyses (n=3).

Two publications, (Huang et al., 2019^111^; Frank et al., 2013^112^) could not be further assessed as results comprised of increased cerebellar volume/positive correlations, and insufficient data was available across cohorts to generate positive CB findings. Three AN publications (Joos et al., 2010^113^; Amianto et al., 2013^114^; D’Agata et al., 2015^115^) included individuals with bulimia nervosa (BN) that contributed to AN findings. As both conditions report with significant diagnostic crossover^116^, these papers were included to form the AN/BN cohort. Due to the limited literature, we were unable to correct for age and gender across datasets.

In summary, 6 OB^117–122^ and 11 AN/BN^113–115,123–128^ papers included coordinates and were selected for final analysis (Participant numbers: nCase-Control=787; nOB=134 vs. nHC=143; nAN=228 & nBN=48 vs. nHC=234) (*Table 1.; Figure 1.*). As an exploratory assessment of condition-specific effects, an AN-only cohort was comprised by excluding three previously included AN papers^113–115^ evaluating those with BN in conjunction with AN (8 papers; nAN=178 vs. nHC=185).

**Figure 1.**
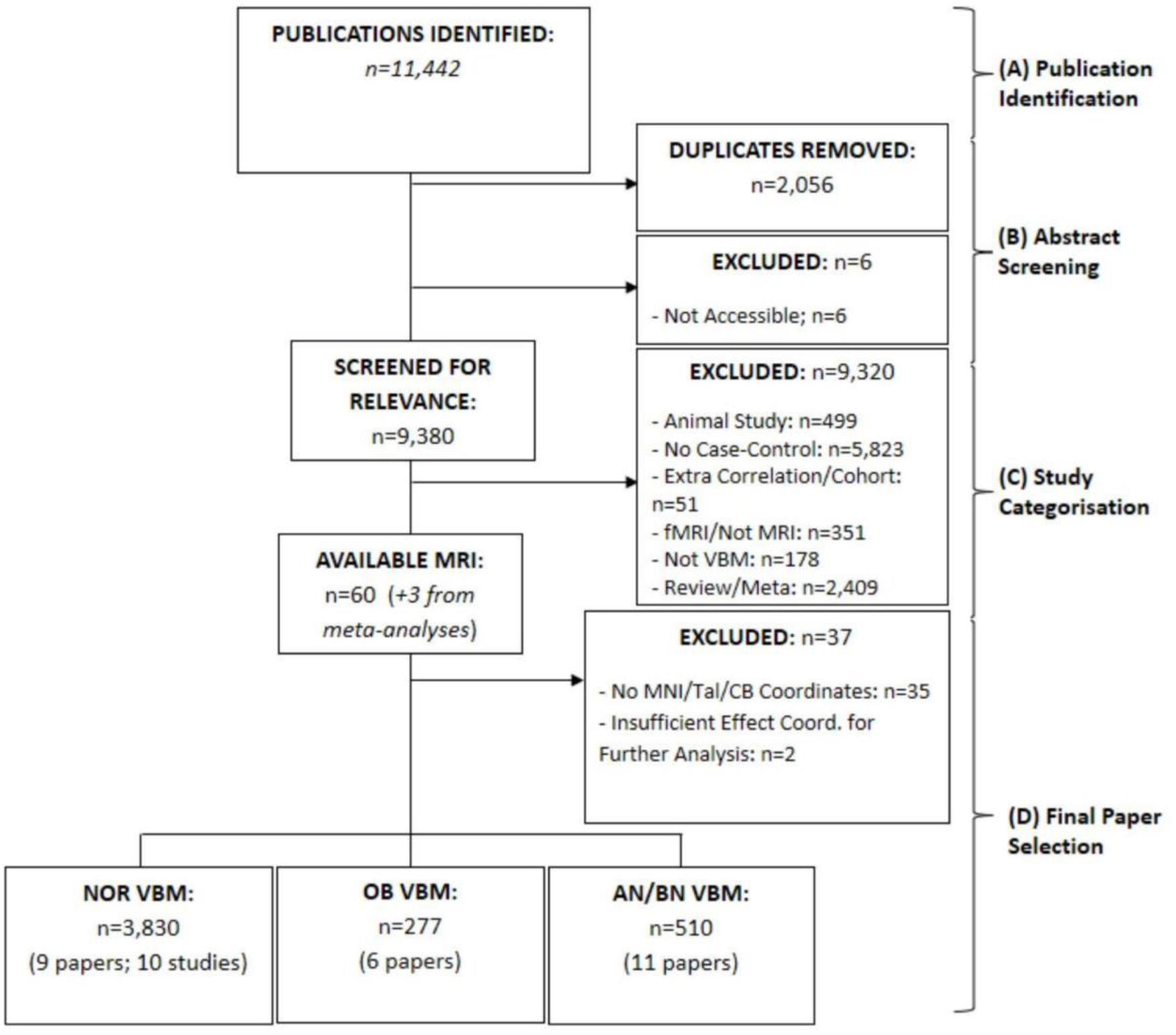
Flowchart depicting the identification (**A**), abstract screening (**B**), categorisation (**C**) and final selection (**D**) of papers. Three papers were also found from meta-analyses (Dommes et al., 2013^120^; Figley et al., 2016^133^; Frank et al., 2013^112^). [**Abbreviations**: AN – Anorexia Nervosa; BN – Bulimia Nervosa; CB – Cerebellum; Coord. - Coordinates; fMRI – Functional Magnetic Resonance Imaging; MNI – Montreal Neurological Institute; MRI – Magnetic Resonance Imaging; NOR – Normative; OB – Obesity; Tal – Talairach; VBM – Voxel-based Morphometry]

**Table 1.**
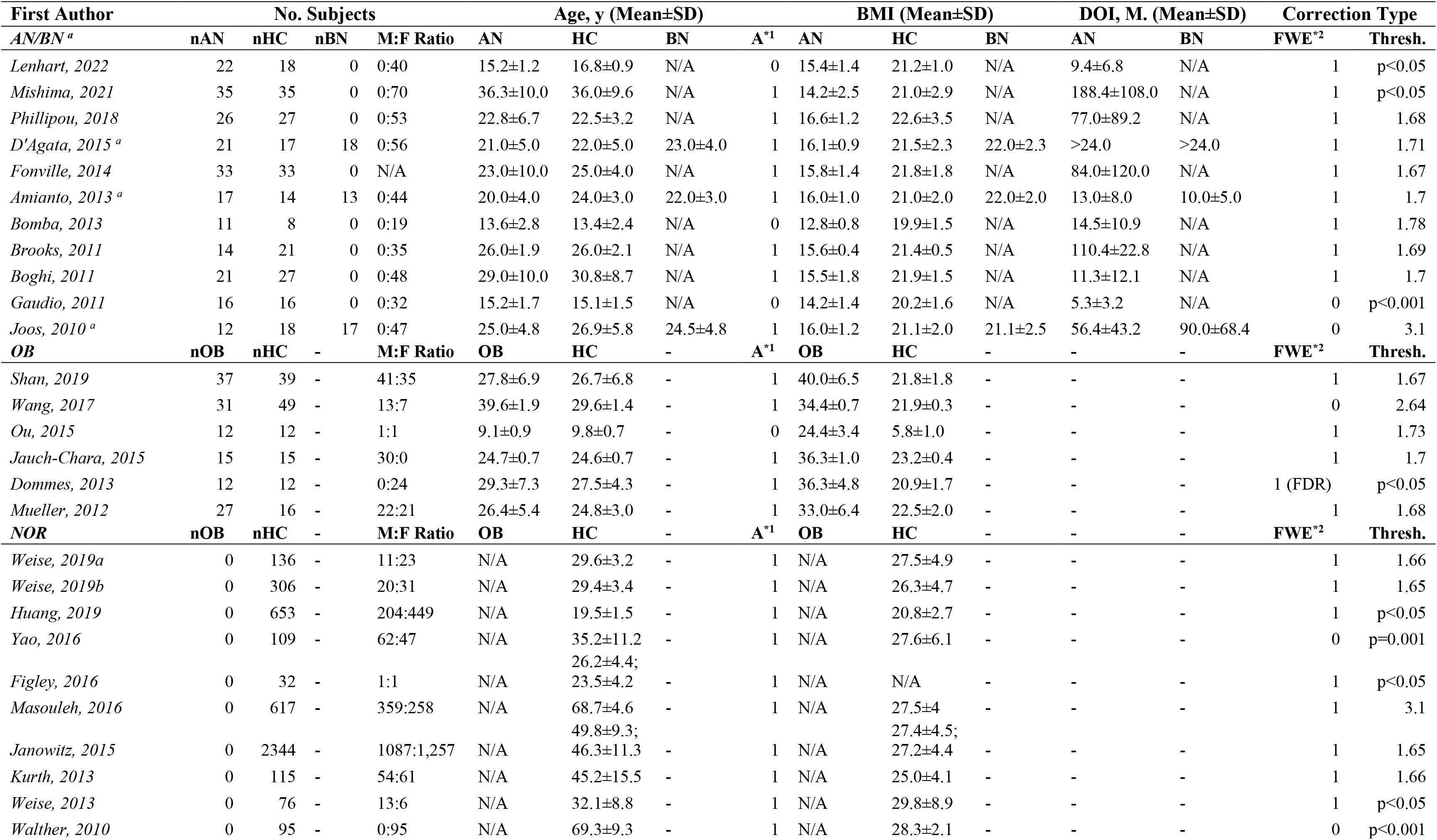
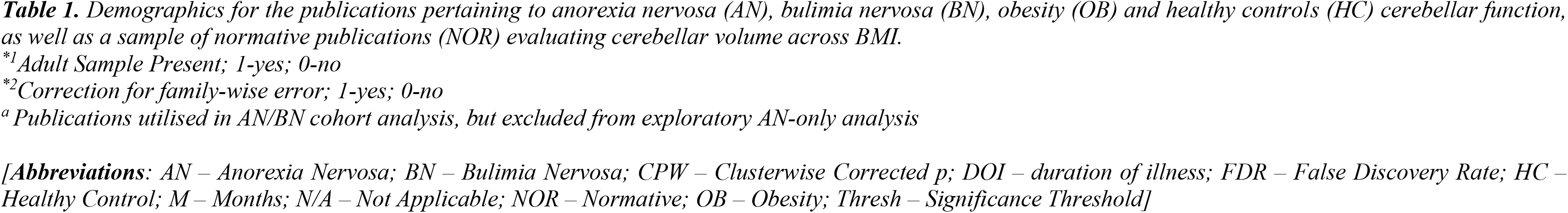
Sociodemographic and statistical data for included studies across AN/BN (n=11), OB (n=6) and NOR (n=10) cohorts.

While conducting the OB literature search, publications were identified evaluating correlations between BMI and GMV in non-clinical, normative (NOR) populations that fell in line with remaining inclusion criteria (n=9 papers; 10 studies). As authors within the NOR subgroup predominantly conducted recruitment using community-based methods and data repositories, participant aggregation generated larger sample sizes than the OB and AN/BN literature. Papers were included in a separate NOR dataset for analysis (participant number=3,830)^129–136^ for a total participant sample size of n=4,617.

### 2.2 Voxel-Based Morphometry and ALE Analysis

Cerebellar coordinates (values depicted in *Table S1)* were respectively incorporated into GingerALE, an Activation Likelihood Estimate (ALE) meta-analysis software using both Talairach and MNI space^137^. GingerALE converted Talairach coordinates to MNI space using the icbm2tal transform. P thresholds for individual analyses were conducted at the whole-brain level and corrected for family-wise error (FWE; 0.05) at 1,000 permutations and set to p<0.01. Firstly, individual observations on respective AN/BN (n=510) and OB (n=277) cohorts were conducted, with a third analysis investigating negative correlations between BMI and cerebellar volume within a NOR data set (n=3,830). As an additional analysis, the AN dataset was re-assessed upon exclusion of publications^113–115^ evaluating volumetric differences in BN (AN-only), but not further investigated for overlap. Condition-respective findings were assessed for overlap via logical overlays and conjunction analyses to visualise combinatorial clusters within AN/BN-OB, AN/BN-NOR, and OB-NOR datasets. Jackknife analyses were conducted to assess robustness of findings via identical re-assessment of cohort data upon one-by-one omission of publications. Additional information regarding cerebellar parcellation, subregion locations, and conjunction analysis thresholding is described within the *Supporting Information*.

## 3.0 Results

### 3.1 Volumetric Reduction in Obesity

Within OB studies, volume of the cerebellum was found to be decreased in the left cerebellar hemisphere, with no bilateral effect. The cluster comprised of 3.58 cm^3^ with a cluster centre of −27, −56, −31. Analysis revealed four peaks of significance partially anteriorly and posteriorly located (*Table 2.*). Specific cerebellar regions affected are Lobule VI, Crus I and the dentate gyrus (*Figure 2.*).

**Table 2.** Coordinates of significance in OB studies (n=6)

Decreased cerebellar GMV in OB cases compared with HCs (OB < HC) including all OB studies (n = 6). Data is presented in order of significance, with the bolded coordinates comprising of the cluster centre.
| Region (OB<HC) | MNI |  |  | Volume (cm<sup>3</sup>) | ALE<br>Score | P | Z |
| --- | --- | --- | --- | --- | --- | --- | --- |
|  | Coordinates |  |  |  |  |  |  |
|  | *x* | *y* | *z* |  |  |  |  |
| *L CB* | -27 | -56 | -31 | 3.58 | 0.00960 | 5.89E-05 | 3.85 |
| *L CB, Anterior L.* | -32 | -56 | -28 |  | 0.00956 | 5.89E-05 | 3.85 |
| *L CB, Anterior L.* | -30 | -60 | -26 |  | 0.00941 | 6.13E-05 | 3.84 |
| *L CB, Posterior L.* | -28 | -54 | -34 |  | 0.00920 | 1.36E-04 | 3.64 |
| *L CB, Posterior L.* | -18 | -54 | -34 |  | 0.00878 | 2.27E-04 | 3.51 |

[**Abbreviations:** ALE – Activation Likelihood Estimation; CB – Cerebellum; HC – Healthy Controls; L – Left; L. – Lobe; MNI – Montreal Neurological Institute; OB – Obesity; R - Right]

**Figure 2.**
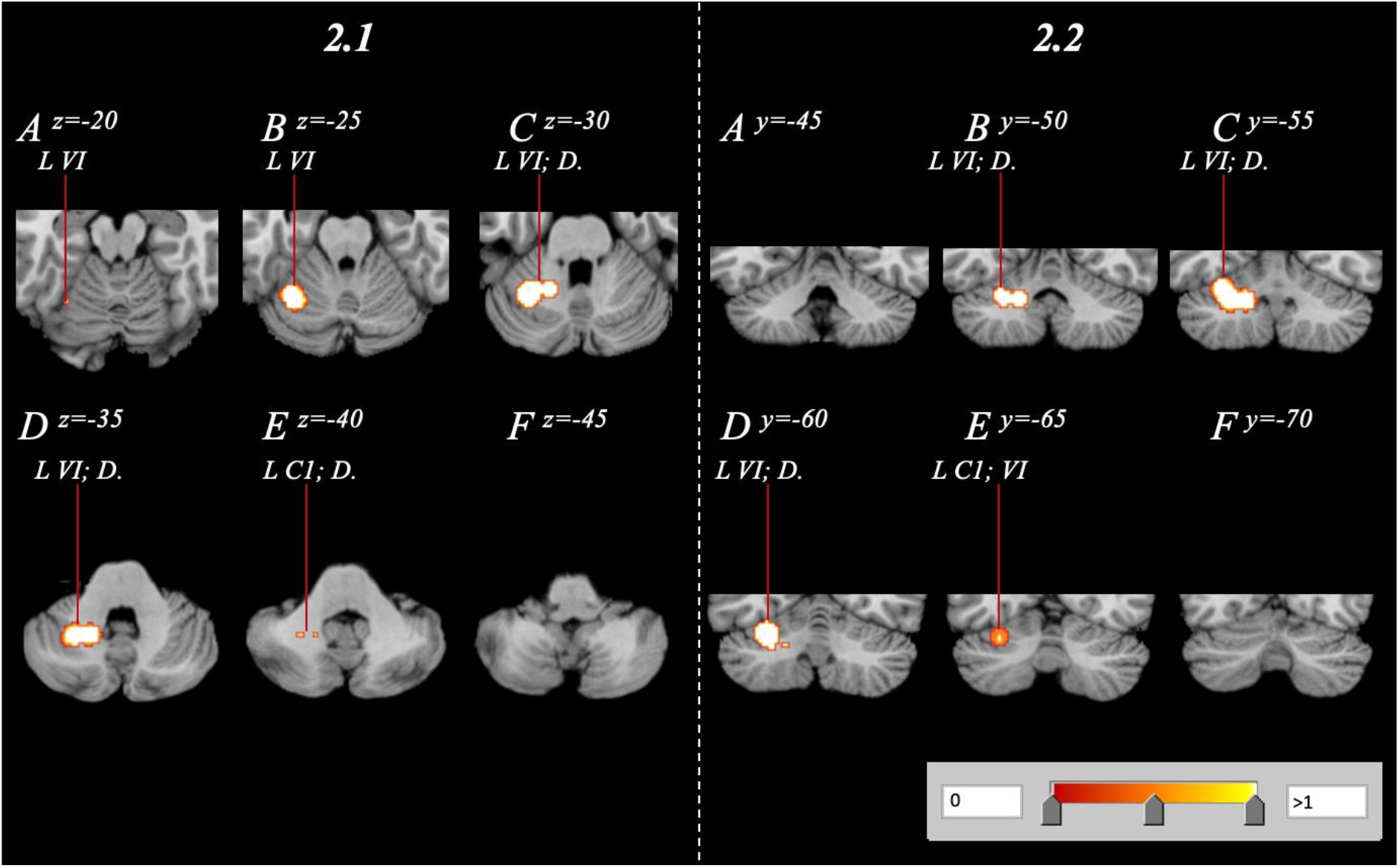
Cerebellar volume reduction of OB subjects relative to HCs, spanning the right cerebellum with a range of z=-20-z=-40 and y=-50-y=-65 in both axial (**2.1A-F**) and coronal (**2.2A-F**) orientations. [**Abbreviations**: C1 – Crus 1; D. – Dentate; L – Left; R – Right; VI – Lobule 6]

### 3.2 Volumetric Reduction in Anorexia Nervosa

The AN/BN GingerALE analysis revealed two significant clusters displaying bilateral reduction of cerebellar volume. The larger cluster was located within the left posterior lobe, contained a volume of 7.09 cm^3^ with a cluster centre of −25, −55, −31. The region contained an additional 7 peaks, with 4 of these occurring in the posterior lobe (*Table 3.*). The smaller cluster comprising of 4.56 cm^3^ was located in the right hemisphere, with 3/5 peaks located in the anterior lobe. Identified subregions included Crus I, the bilateral Lobule IV, and the bilateral dentate (*Figure 3*.). Contrasting AN/BN findings, the exploratory AN-only GingerALE analysis revealed no significant clusters displaying volumetric reduction within the cerebellum.

**Table 3.**
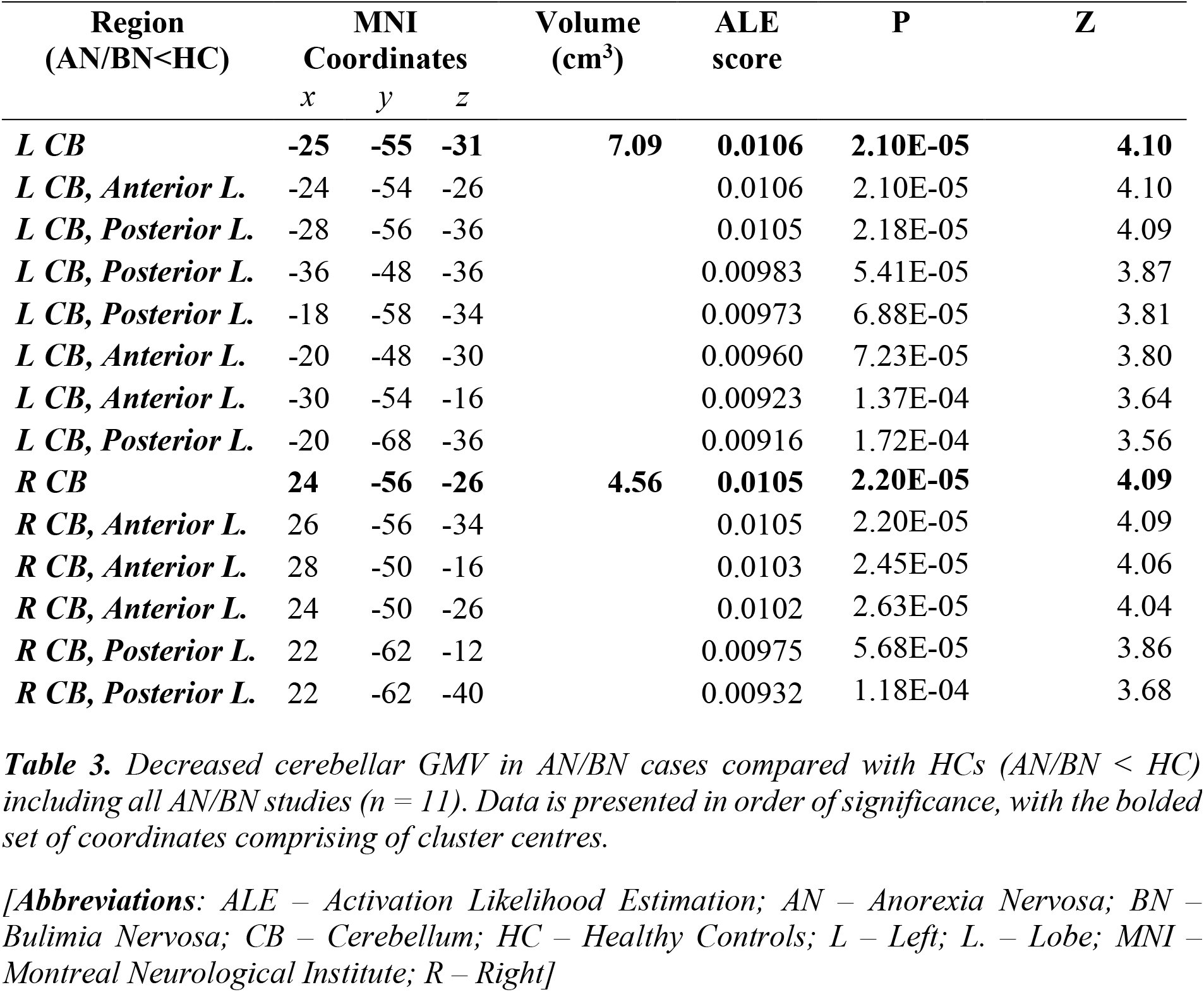
Coordinates of significance in AN/BN studies (n=11)

Decreased cerebellar GMV in AN/BN cases compared with HCs (AN/BN < HC) including all AN/BN studies (n = 11). Data is presented in order of significance, with the bolded set of coordinates comprising of cluster centres.
| Region<br>(AN/BN<HC) | MNI<br>Coordinates |  |  | Volume<br>(cm<sup>3</sup>) | ALE<br>score | P | Z |
| --- | --- | --- | --- | --- | --- | --- | --- |
|  | x | y | z |  |  |  |  |
| **L CB** | **-25** | **-55** | **-31** | **7.09** | **0.0106** | **2.10E-05** | **4.10** |
| *L CB, Anterior L.* | -24 | -54 | -26 |  | 0.0106 | 2.10E-05 | 4.10 |
| *L CB, Posterior L.* | -28 | -56 | -36 |  | 0.0105 | 2.18E-05 | 4.09 |
| *L CB, Posterior L.* | -36 | -48 | -36 |  | 0.00983 | 5.41E-05 | 3.87 |
| *L CB, Posterior L.* | -18 | -58 | -34 |  | 0.00973 | 6.88E-05 | 3.81 |
| *L CB, Anterior L.* | -20 | -48 | -30 |  | 0.00960 | 7.23E-05 | 3.80 |
| *L CB, Anterior L.* | -30 | -54 | -16 |  | 0.00923 | 1.37E-04 | 3.64 |
| *L CB, Posterior L.* | -20 | -68 | -36 |  | 0.00916 | 1.72E-04 | 3.56 |
| **R CB** | **24** | **-56** | **-26** | **4.56** | **0.0105** | **2.20E-05** | **4.09** |
| *R CB, Anterior L.* | 26 | -56 | -34 |  | 0.0105 | 2.20E-05 | 4.09 |
| *R CB, Anterior L.* | 28 | -50 | -16 |  | 0.0103 | 2.45E-05 | 4.06 |
| *R CB, Anterior L.* | 24 | -50 | -26 |  | 0.0102 | 2.63E-05 | 4.04 |
| *R CB, Posterior L.* | 22 | -62 | -12 |  | 0.00975 | 5.68E-05 | 3.86 |
| *R CB, Posterior L.* | 22 | -62 | -40 |  | 0.00932 | 1.18E-04 | 3.68 |

[Abbreviations: ALE – Activation Likelihood Estimation; AN – Anorexia Nervosa; BN – Bulimia Nervosa; CB – Cerebellum; HC – Healthy Controls; L – Left; L. – Lobe; MNI – Montreal Neurological Institute; R – Right]

**Figure 3.**
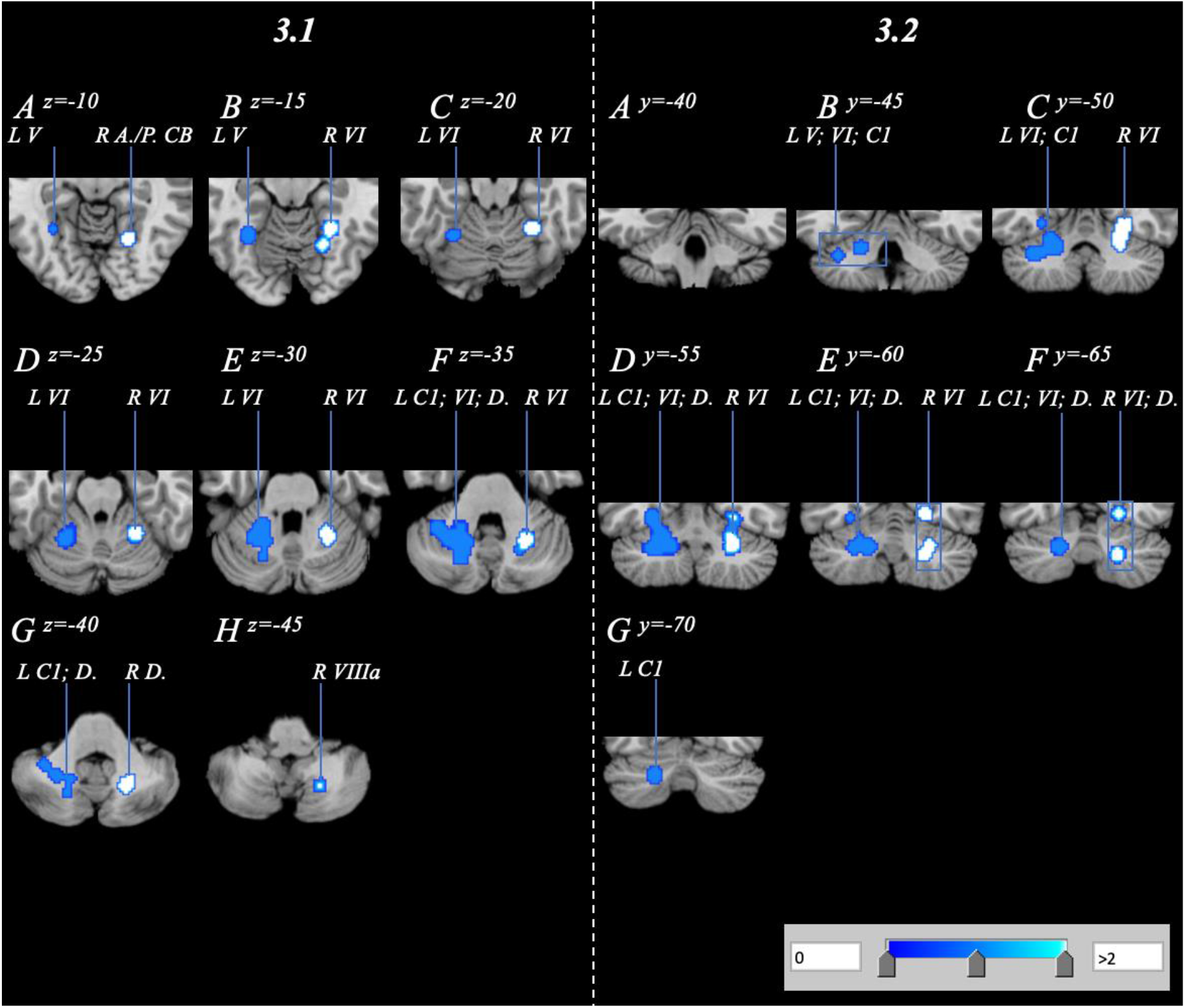
Cerebellar volume reduction of AN/BN subjects relative to HCs, spanning the bilateral cerebellum with a range of z=-10-z=-45 and y=-45-y=-70 in axial (**3.1A-H**) and coronal (**3.2A-G**) orientations. [**Abbreviations**: A. – Anterior; C1 – Crus 1; CB – Cerebellum; D. – Dentate; L – Left; P. – Posterior; R – Right; VI – Lobule 6; VIIIa – lobule 8a]

### 3.3 Normative Analysis: Cerebellum GMV vs. BMI

Analysis of the NOR dataset evaluating correlations between volumetric measures and BMI revealed a singular posterior cluster where cerebellum GMV negatively correlated with BMI. The cluster had a volume of 8.02 cm^3^ with a cluster centre at 30, −71, −32 (*Table 4.*; *Figure 4.*). The cluster comprised of 7 peaks, all located within the posterior lobe affecting regions such as right Crus I/II, Lobule VI, Lobule VIIb and the dentate.

**Table 4.** Clusters associated with increased BMI in NOR populations (n=9/10)

Decreased cerebellar GMV in NOR populations when correlated with BMI and weight class ( $n = 9$ papers, 10 studies). Data is presented in order of significance, with the bolded set of coordinates representing the cluster centre.
| (NOR; CB GMV<br>vs. BMI) | MNI<br>Coordinates |  |  | Volume (cm<sup>3</sup>) | ALE score | P | Z |
| --- | --- | --- | --- | --- | --- | --- | --- |
|  | x | y | z |  |  |  |  |
| **R CB** | **30** | **-71** | **-32** | **8.02** | **0.0224** | **1.39E-08** | **5.56** |
| *R CB, Posterior L.* | 28 | -70 | -42 |  | 0.0224 | 1.39E-08 | 5.56 |
| *R CB, Posterior L.* | 44 | -72 | -30 |  | 0.0207 | 5.54E-08 | 5.31 |
| *R CB, Posterior L.* | 18 | -66 | -30 |  | 0.0189 | 4.14E-07 | 4.93 |
| *R CB, Posterior L.* | 28 | -80 | -24 |  | 0.0138 | 1.50E-05 | 4.17 |
| *R CB, Posterior L.* | 34 | -78 | -28 |  | 0.0133 | 1.91E-05 | 4.12 |
| *L CB, Posterior L.* | 36 | -72 | -18 |  | 0.011 | 6.86E-05 | 3.81 |

[**Abbreviations:** ALE – Activation Likelihood Estimation; CB – Cerebellum; L – Left; L. – Lobe; GMV – Grey Matter Volume; MNI – Montreal Neurological Institute; NOR – Normative; R – Right]

**Figure 4.**
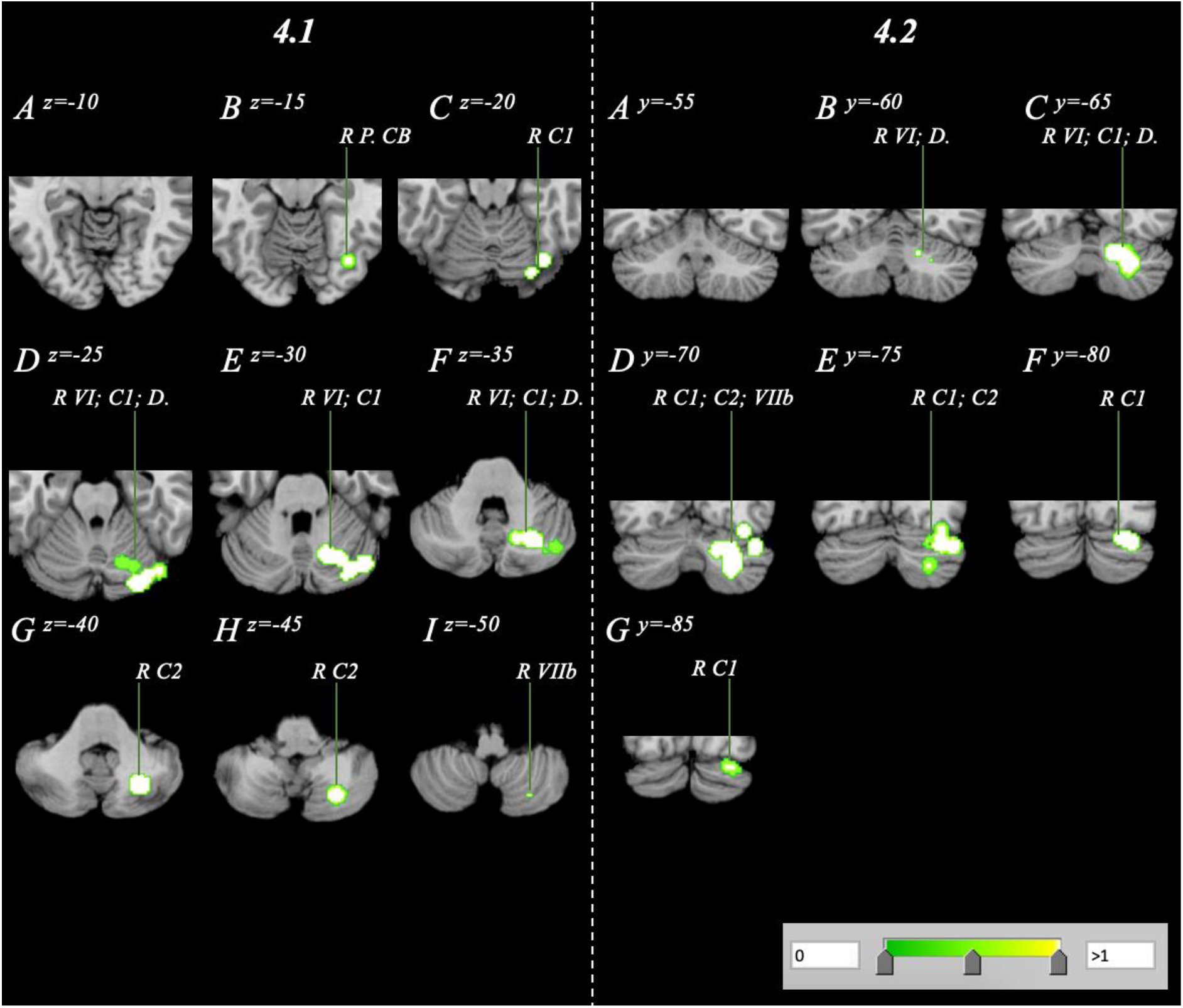
Volume reduction within a normative sample analysis spanning the right cerebellum with a range of z=-15-z=-50 and y=60-y=-85 in axial (**4.1A-I**) and coronal (**4.2A-G**) orientations. [**Abbreviations**: C1 – Crus 1; C2 – Crus 2; D. – Dentate; L – Left; P. – Posterior; R – Right; VI – Lobule 6; VIIb – Lobule 7b; VIIIa/b – Lobule 8a/b]

### 3.4 Pooled Volumetric Reduction and Conjunction Analyses

Pooling combinations of data (AN/BN-OB, AN/BN-NOR, OB-NOR) showed that cerebellar volume reduction both significantly overlapped across cohorts but were also largely distinct according to bodyweight condition as well as across BMI (*Figure S2.)*. OB and NOR clusters were exclusive to the left and right hemispheres respectively, while AN/BN clusters were bilaterally located. Logical overlays on MANGO delineated significant inter-condition overlap, which prompted three conjunction analyses (*Figure 5.*). The AN/BN-OB conjunction analysis revealed a large region of overlap. The cluster was located in the left Lobule VI with a volume of 0.214 cm^3^ and centre coordinates of −24, −55, −33 (*Figure 6.1.; Table 5.*). Conjunction analyses between clinical cohorts and the NOR cohort conducting correlational analyses between volume and BMI revealed additional findings. The AN/BN-NOR conjunction analysis revealed a unilateral overlap of structural reduction. The cluster was 0.0296 cm^3^ with a cluster centre of 23, −64, −39, and primarily located within right Lobule VI, but also affected the surrounding Crus II and Dentate Nucleus regions (*Figure 6.2.; Table 5.)*. No overlap or combinatorial clusters falling within our significance threshold were found in the OB-NOR conjunction analysis.

**Figure 5.**
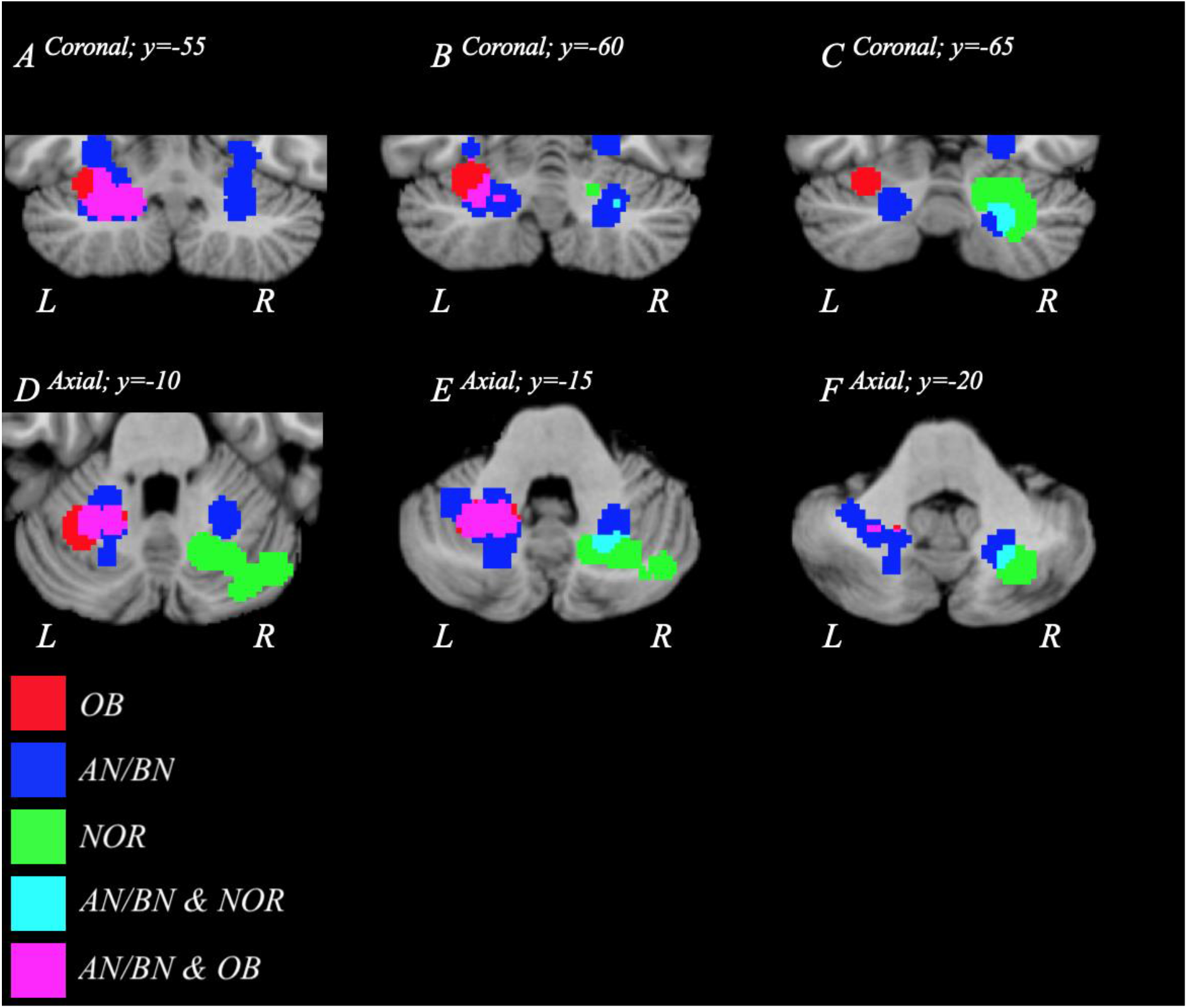
FWE-corrected overlay clusters visualising decreases in volume from AN/BN (blue) OB (red) and NOR (green) data. Regions of the cerebellum where AN/BN and OB data overlap are shown in purple, while an overlap in AN/BN and NOR data are displayed in cyan. No overlap between OB and NOR data was present. Figures 5A-C and D-F depict the logical overlays in axial and coronal orientations respectively. [**Abbreviations**: AN – Anorexia Nervosa; BN – Bulimia Nervosa; L – Left; NOR – Normative; OB – Obesity; R – Right]

**Figure 6.**
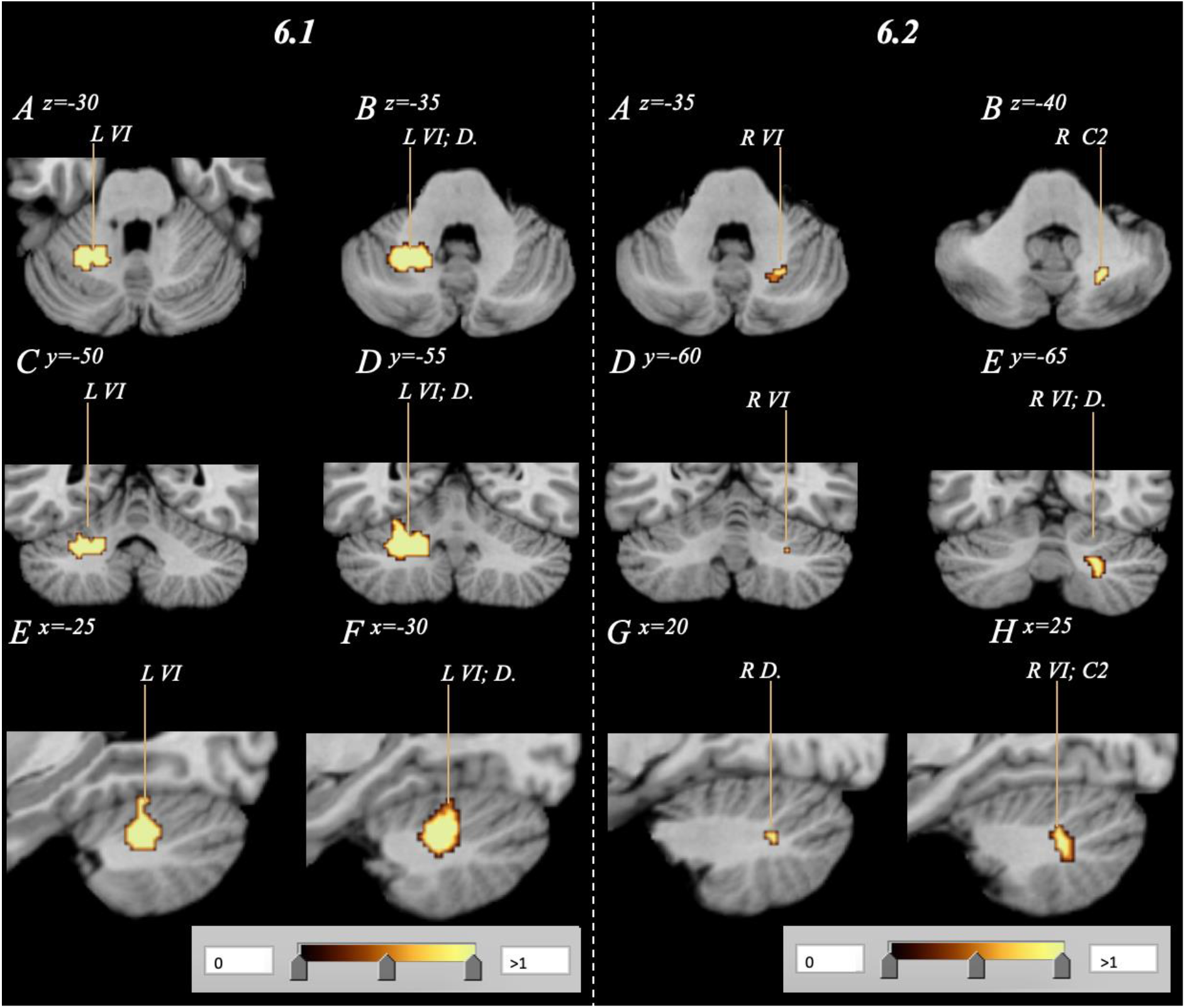
Subsequent conjunction analysis combining respective AN/BN and OB data (**6.1**) as well as AN/BN and NOR data (**6.2**) to visualise affected regions of overlap. Analyses confirm an overlap in volumetric decrease in both AN/BN and OB subjects, as well as AN/BN and NOR subjects primarily within Lobule VI (p<0.05). Subfigures 6A/B, C/D and D/E depict the combinatorial clusters in axial, coronal and saggital orientations respectively. No overlap between OB and NOR data was identified. [**Abbreviations**: AN – Anorexia Nervosa; BN – Bulimia Nervosa; C2 – Crus 2; D. – Dentate; L – Left; NOR – Normative; OB – Obesity; R – Right; VI – Lobule 6]

**Table 5.**
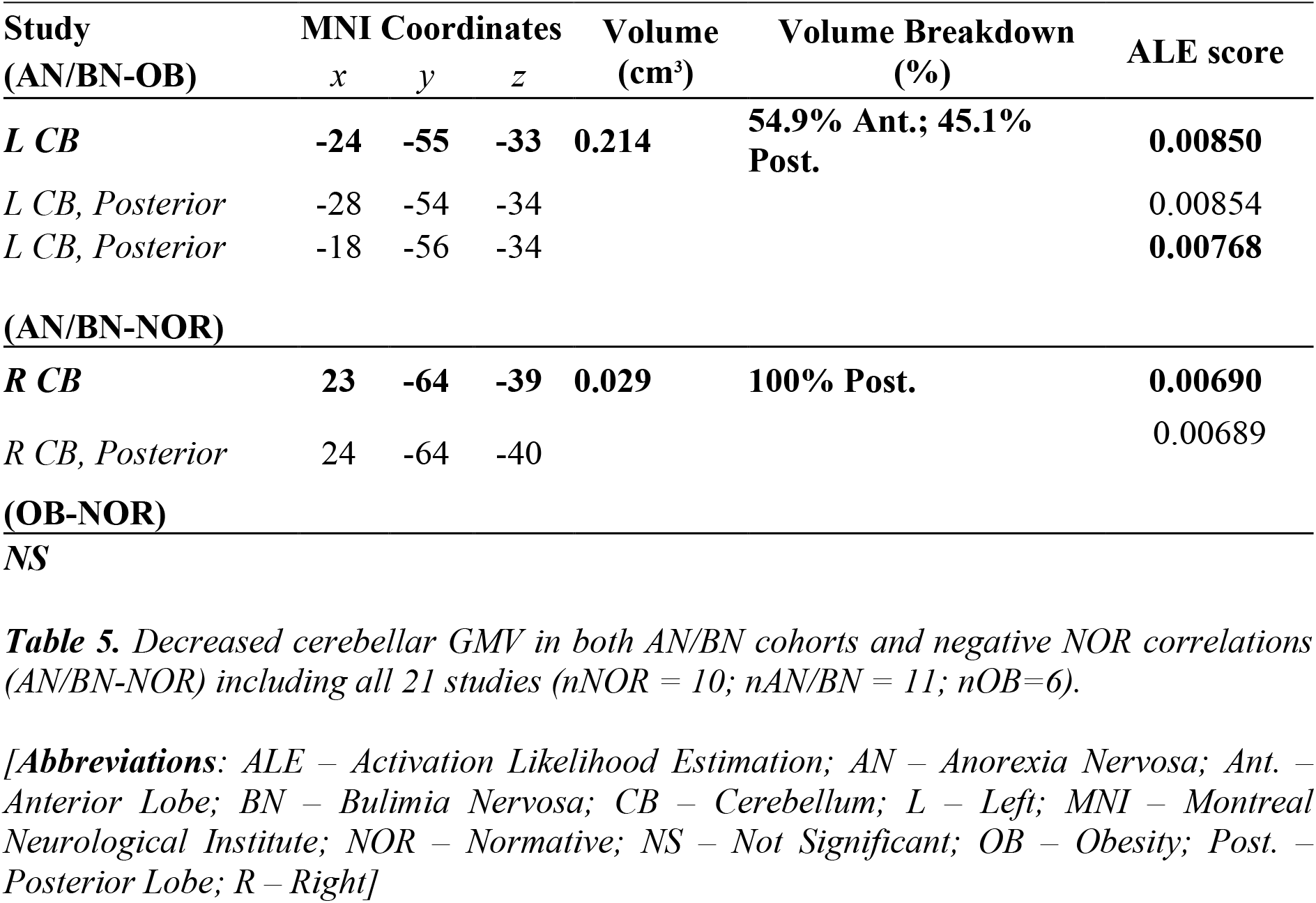
Conjunction analysis clusters of significance in AN/BN, NOR and OB cohorts.

Decreased cerebellar GMV in both AN/BN cohorts and negative NOR correlations (AN/BN-NOR) including all 21 studies (*n*NOR = 10; *n*AN/BN = 11; *n*OB=6).
| Study<br>(AN/BN-OB) | MNI Coordinates |  |  | Volume<br>(cm<sup>3</sup>) | Volume Breakdown<br>(%) | ALE score |
| --- | --- | --- | --- | --- | --- | --- |
|  | *x* | *y* | *z* |  |  |  |
| ***L CB*** | **-24** | **-55** | **-33** | **0.214** | **54.9% Ant.; 45.1% Post.** | **0.00850** |
| *L CB, Posterior* | -28 | -54 | -34 |  |  | 0.00854 |
| *L CB, Posterior* | -18 | -56 | -34 |  |  | **0.00768** |
| **(AN/BN-NOR)** |  |  |  |  |  |  |
| ***R CB*** | **23** | **-64** | **-39** | **0.029** | **100% Post.** | **0.00690** |
| *R CB, Posterior* | 24 | -64 | -40 |  |  | 0.00689 |
| **(OB-NOR)** |  |  |  |  |  |  |
| **NS** |  |  |  |  |  |  |

[**Abbreviations:** ALE – Activation Likelihood Estimation; AN – Anorexia Nervosa; Ant. – Anterior Lobe; BN – Bulimia Nervosa; CB – Cerebellum; L – Left; MNI – Montreal Neurological Institute; NOR – Normative; NS – Not Significant; OB – Obesity; Post. – Posterior Lobe; R – Right]

### 3.5 Sensitivity Analyses

Sensitivity analyses were conducted on all available cohort datasets (*Table S2*.). One-by-one omission of publications identified heterogeneity across OB findings, in which the left posterior and anterior cerebellar clusters were present in 3/6 analyses. Alternatively, NOR and AN/BN findings were robust, with the right posterior cerebellar cluster demonstrating repeatability in 10/10 NOR analyses, as well as left and right AN/BN cerebellar clusters appearing in 11/11 and 9/11 analyses respectively. While the AN-only cohort did not report findings, the jackknife analysis identified the left posterior CB as reduced upon removal of Lenhart et al. (2022)^126^, Phillipou et al. (2018)^125^, Fonville et al. (2014)^124^ and Bomba et al. (2013)^123^.

## 4.0 Discussion

### 4.1 Cerebellar Differences: OB vs. AN/BN vs. HC

Compiling data from OB and AN publications, as well as correlations between BMI and brain volume, confirms cerebellar structure to be significantly decreased at both extremes of the spectrum of bodyweight disorders. Regions Crus I and Lobule VI were most consistently and significantly affected, identifying two cerebellar regions with respective executive and sensorimotor functionality associated with conditions of dysregulated BMI. In addition, findings indicate both condition-specific differences as well as significant overlap in cerebellar GMV reduction between AN/BN, OB and NOR cohorts. Reduced GMV in Lobule VI, Crus I and the dentate nucleus was present in all cohorts, but differed according to hemisphere. In the NOR population, affected regions correlating with increased BMI overlapped with cerebellar reduction seen in AN/BN, but not the OB analysis. Differential hemispheric involvement of two regions that reportedly comprise distinct cortico-cerebellar circuits leads us to suggest that multiple cerebellar regions are reliably and consistently involved across states of appetite dysfunction. Understanding of these regional functions may provide insight as to how the cerebellum may differentially contribute to the dysregulation of body-weight.

Crus I, structurally reduced in all cohorts, is reported to play predominant executive-, memory- and some emotional-related functions. Crus I and II contain specific functional localisation that corresponds to cerebral cortical zones^37,39,138–140,143^ and falls within the executive control network (ECN)^37–39^. In typical populations, this network plays roles in executive function, spatial attention^144^ and verbal working memory^39,140–143^, as well as goal-directed behaviour via communication with the hippocampus^142^. Recently, Crus II is reported to serve emotional self-experience and social mentalising^145^, expanding emotional roles played by posterior cerebellar lobes.

Functional hemispheric differences reported in Crus I may explain how the cerebellum is implicated, but likely plays distinct roles in cases of pathological under- and over-eating. Bilateral cerebellar reduction has previously been associated in those with AN^146^. Despite no cluster-based findings within the exploratory AN-only cohort, Crus I reduction within the AN/BN cohort may be associated with general executive and attentional deficits reported in undereating^147^, such as body-image disturbance and behavioural rigidity towards food. Alternatively, reduction of Crus I within this cohort may be moreso associated with BN-specific symptomatology, such as altered traits of impulsivity or LOC eating. Additionally, volumetric reduction of Crus II was only identified within the NOR and AN/BN findings, suggesting socio-emotional functions of Crus II could play a less prominent role in those with OB. Crus I reduction was specific to the left hemisphere in the OB cohort, which interestingly overlaps with previous unilateral cerebellar PSE^105^ findings as well as regions activated in spatial attention^144^ and social processing tasks^143^. Thus, cerebellar volume reduction in cases of over-eating or OB seen in this meta-analysis may suggest altered measures of food-related attention, impulsivity, working memory or loss of control, aspects characteristic of OB^148–153^. However, sensitivity analyses reported heterogeneity amongst OB study findings, likely due lacking study from limited literature. Further region-of-interest structural studies as well as resting-state and fMRI tasks relating to impulsivity, working memory and decision making would aid in furthering condition-specific interpretations from this work.

Within NOR populations, reduction of Crus I was associated with increased BMI, but reduction was unilateral to the right hemisphere and included reduction of Crus II, suggesting differential mechanisms underlying non-pathological weight gain in comparison to states of OB. While it is important to note the differential study power between cohorts, with NOR findings derived from a significantly larger sample size than OB findings, findings nonetheless corroborate that the cerebellum is differentially associated with non-pathological weight gain.

Lobule VI consistently identified as structurally reduced within all cohorts, and its known functional distinction from Crus I furthers the concept that more than one region of the cerebellum participates in aspects of the dysregulation of body-weight. Lobule VI has recently been recognised as serving several cognitive and emotional functions, and plays roles in the salience^37,39,154–156^, somatomotor, ventral attention and visual networks^145^. Roles of the salience network, predominantly represented via Lobule VI, involve autonomic and interoceptive processing in response to various forms of salience such as emotion, reward, and homeostatic regulation^157–159^. Tractography studies demonstrate connections between the motor cortex, Lobule VI and dentate nucleus^160,161^, and fMRI studies report consistent Lobule VI activity in emotional, social and environmental learning tasks^162–164^. Computational models also support that circuits implicated Lobule VI are important for Pavlovian conditioning and emotional learning^165–169^, including perceptual inferences such as startle responses and spontaneous behaviour^170^. Such mechanisms may contribute to AN or BN via development of food aversion or negative food/weight associations which tend to exacerbate low dietary intake or restriction. Alternatively, food may be conditioned as an excessively rewarding or positive stimulus, which may be associated with overeating or LOC consumption.

Similar to Crus I findings, differential hemispheric reduction of Lobule VI across cohorts suggests differential cerebellar involvement. While the exploratory AN-only analysis identified no clusters of interest, the AN/BN analysis identified bilateral reduction of Lobule VI. Those with AN have previously demonstrated reduced bilateral volume of the mid-posterior cerebellum, which is suggested to contribute towards characteristic AN symptoms such as food aversion^146^, but further functional studies focusing on the cerebellum are warranted to further understand this relationship, as well as additional studies to specify between AN and BN findings. Reduction of Lobule VI in OB was restricted to the left hemisphere, which contains regions specific to social processing tasks^143^. In contrast to the OB cohort, reduction of Lobule VI volume within the NOR population was specific to the right hemisphere. While there is currently no specific ascribed role to the right Lobule VI, differing hemisphericity of effect between OB and NOR cohorts similarly suggests differential cerebellar participation between pathological/non-pathological overeating.

Lastly, reduction of the dentate nucleus was seen in all cohorts. This region is highly connected to the hypothalamus and thought to interact with the lateral hypothalamic area, ventromedial nucleus, dorsomedial nucleus, and paraventricular nucleus to modulate forelimb movements in food grasping behaviour^171^. As with previous findings, the recruitment of the dentate nucleus within multiple cohorts not only suggests cerebellar implication in both non-pathological and clinical states of body-weight dysregulation, which likely functionally differs due to associations with opposing hemispheres, but also that multiple cerebellar regions are associated with the dysregulation of bodyweight and BMI.

### 4.2 Cerebellar Overlap: OB vs. AN/BN vs. HC

Volumetric overlap among cohorts revealed that cases of dysregulated appetite across the BMI-spectrum exhibit significant reduction in similar cerebellar regions, which have been known for distinct sensorimotor (Lobule VI) and executive (Crus I/II) roles and suggest multimodal cerebellar association across the BMI spectrum. Structural reduction in cases of AN/BN and OB, as well as within NOR populations assessing cerebellar volume alongside increased BMI, was predominant to Lobule VI and Crus I. Findings relating to non-pathological BMI increase via the NOR cohort as well as the AN/BN cohort overlapped within the right Lobule VI, Crus II and dentate nucleus. Alternatively, Lobule VI and Crus I were both reduced in OB and NOR datasets, but difference of effect according to hemisphere meant no overlap in volume reduction was identified. The implication of similar cerebellar regions yet differing hemisphericity of effect again suggests that while these cerebellar regions participate in differing forms of body-weight states, the main sources of variation among healthy controls may be quite different to those driving differences in clinical conditions.

### 4.3 Limitations

#### 4.3.1 Number of Studies

Our review was limited by low study numbers used for some individual analyses, and we were unable to provide sufficient information on race/ethnicity and socioeconomic status. Availability of data also prevented further evaluation into sex, age, hormonal differences and brain inflammation values across participants. Due to such limitations, we confined our analysis to changes in GMV. Future research would benefit from utilising more specified imaging technology catered to WMV alterations, such as diffusion tensor- and weighted-imaging.

Similarly, while there is no definitive instruction on the minimum number of studies needed to conduct an ALE meta-analysis, the GingerALE (2.0) manual mentions that a minimum of 20-30 coordinates per experiment, as seen in our analyses, is sufficient to produce significant and valid clusters for simple paradigms. Later on, Eickhoff *et al*. (2016)^172^ recommended approximately 15-17 studies for reliability of analysis results, which were not present in our study. While cohort-specific sensitivity analyses were implemented to attempt to account for heterogeneity across findings, little can be done to correct or adjust for small sample sizes via GingerALE. A method of counteracting potentially unreliable results from future smaller studies would be to analyse small-cohort findings alongside larger datasets, or to incorporate quantifications of bias from small sample sizes (i.e., Egger test) within GingerALE software utility.

#### 4.3.2 Assumptions

The research question posed by this review – to determine if areas of the cerebellum are involved in different disorders of appetite control were the same or distinct, was driven by a review of the literature providing extensive evidence that the cerebellum is implicated in disorders of appetite control (see Introduction). Therefore, our study is based on this assumption and does not independently verify it. This is because we only reviewed studies that included cerebellum findings rather than all MRI studies of eating disorders. Additionally, both AN/BN and OB are associated with elevated inflammation within the brain^173,174^, which may influence findings related to volumetric reduction of the cerebellum. Future studies would benefit from including brain inflammation factors within analysis.

#### 4.3.3 Causality and Direction of Effects

Further, we are unable to verify the direction of causation in found associations. A common limitation in neuroimaging studies is their capacity to determine causality. The studies reviewed here are only able to demonstrate associations between volume abnormalities and condition, and it is unclear whether reduced volumes are a cause or subsequent effect.

#### 4.3.4 Bulimia *Nervosa*

Although we excluded the majority of BN patients, approximately 20% of clinical AN data consisted of those with BN which we were unable to remove. As there were insufficient BN papers (n=3) to conduct individualised cohort analyses for AN and BN, datasets were thus pooled into one AN/BN cohort. While there is a significant overlap in comorbidity and symptomatology between those with AN and BN^116^, including this group may have resulted in volume reduction not linked to states relating to those with AN, and findings from this meta-analysis are unable to distinguish between those with mixed AN/BN pathology and those with AN alone. We attempted to mitigate limitations regarding AN/BN etiological heterogeneity by conducting an exploratory assessment with AN-only publications. Nevertheless, our study still focuses on reduced cerebellar volume in eating disorders, which is clearly the case for bulimic individuals.

## Conclusion

While theories proposing cerebellar functions in emotional, cognition and conditioning behaviours are now receiving wide acceptance, a role in weight and appetite is rarely discussed. In this review, we found utility in exploring cerebellar differences and similarities in states at opposite ends of the bodyweight dimension, and collated structural evidence from many sources. Altogether, results of our ALE analyses support the concept that the cerebellum is associated with states dysregulated appetite, eating disorders and bodyweight, in both pathological and non-pathological states. We found that while cerebellar associations with bodyweight issues recruited similar regions, hemispherical effects differed according to the type of appetitive condition, suggesting different cerebellar circuitry contributing to eating behaviour between non-clinical and pathological populations. Such findings add to a body of evidence theorising cerebellar participation in appetite- and feeding-related domains. Utilising our knowledge of the emotional and cognitive functions of the cerebellum will assist in identifying novel remedial approaches to manage disorders of appetite-regulation as well as increase efficacy of interventions provided to clinical populations.

## Data Availability

Data produced in this study are available upon reasonable request to the corresponding author.

---

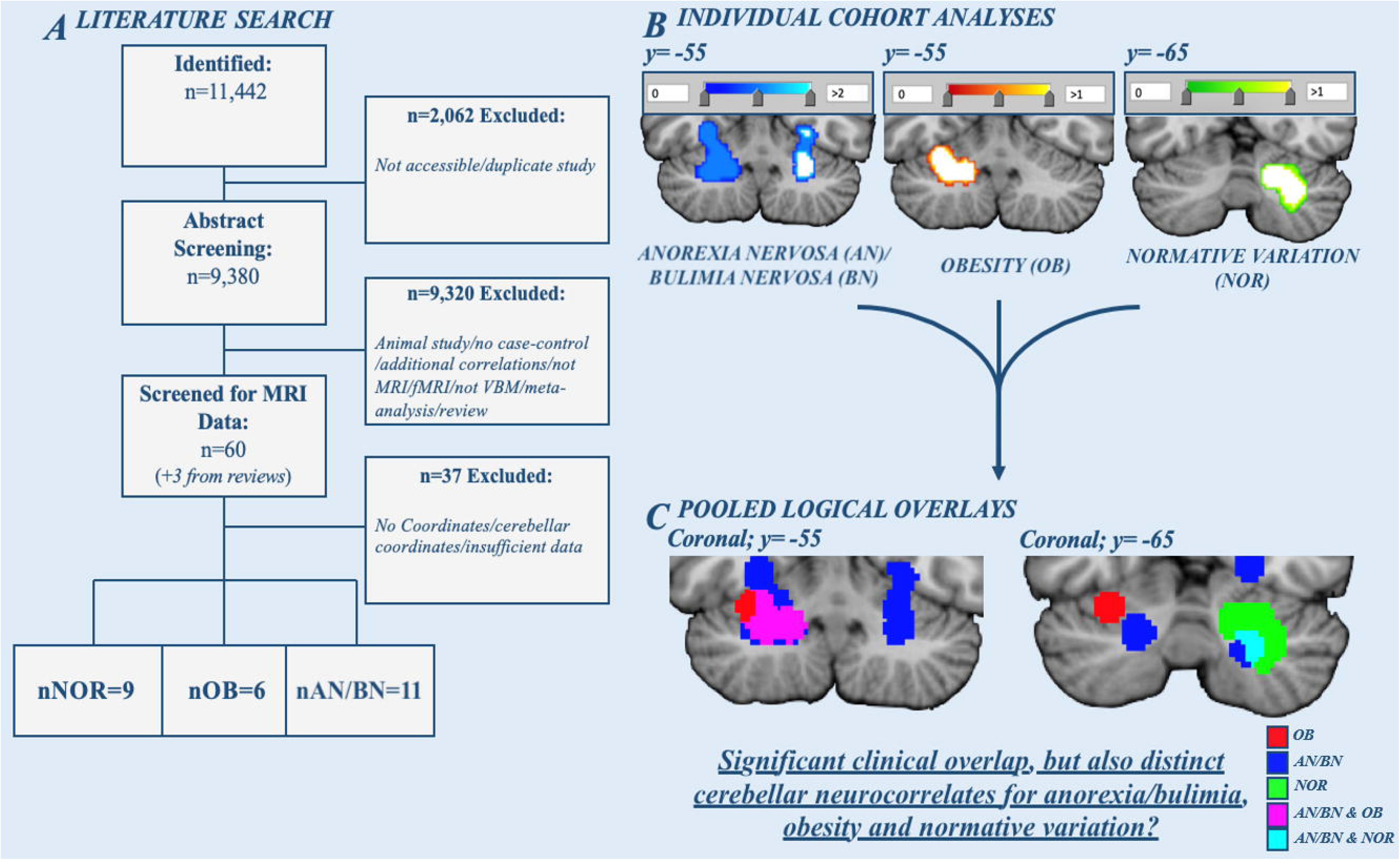

