## Supporting Information for "The Cerebellum Plays More Than One Role in the Dysregulation of Appetite: Review of Structural Evidence from Typical and Eating Disorder Populations"

**Authors:** Michelle Sader[1]*, Gordon D. Waiter[1], Justin H. G. Williams[1,2,3]

[1]Biomedical Imaging Centre, University of Aberdeen, United Kingdom

[2]School of Medicine, Griffith University, Queensland, Australia

[3]Gold Coast Mental Health and Specialist Services, Gold Coast, Queensland, Australia

---

---

#### 1.0 – Introduction

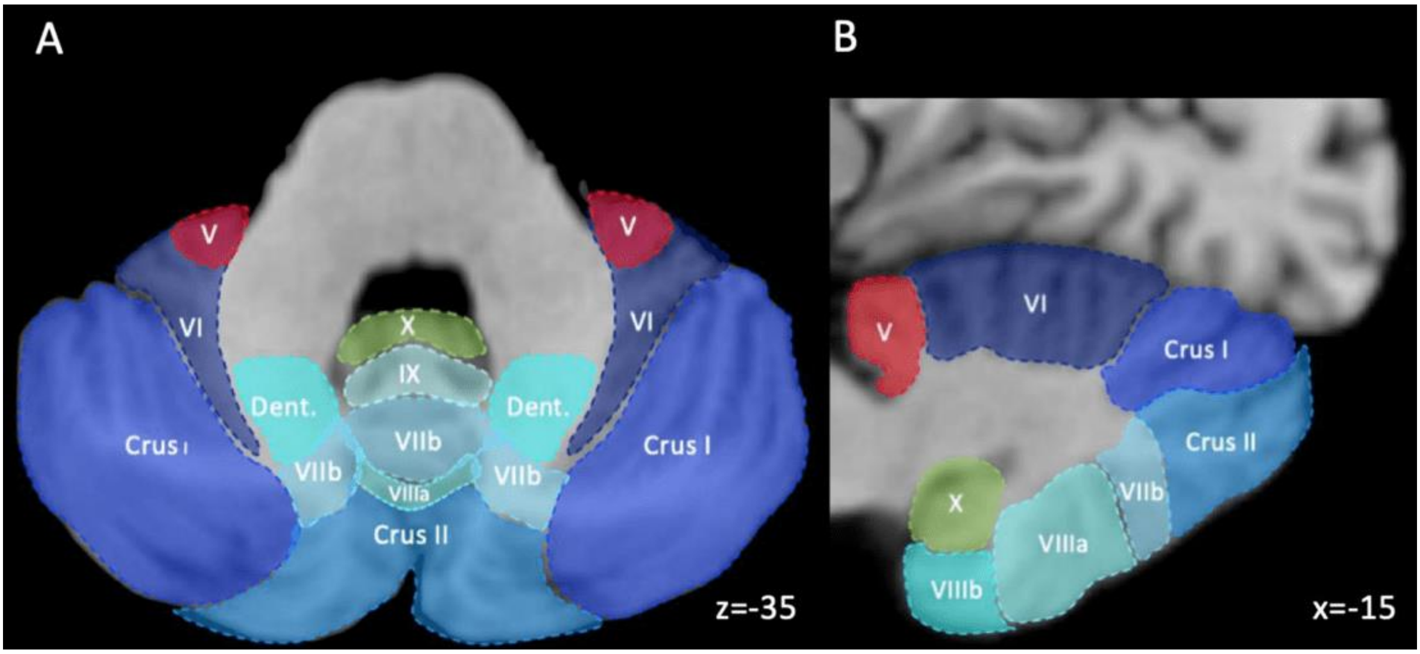


***Figure S1.*** *Schematic of the human cerebellum in an axial (**A**) and saggital (**B**) orientation, depicting the anterior (red), posterior (blue) and flocculonodular (green) lobes. Cerebellar subregions visualised under the following MNI coordinates have been demarked by the SUIT cerebellar atlas[1].*

*[**Abbreviations**: Dent. – dentate gyrus; V – lobule 5; VI – lobule 6; VIIb – lobule 7b; VIIIa – lobule 8a; IX – lobule 9; X – lobule 10]*

### 2.0 – Methodology

#### *2.2 – Voxel-Based Morphometry and ALE Analysis*

Each selected publication contained sets of coordinates indicating difference in cerebellar volume. While selected literature contained whole-brain analyses, only coordinates pertaining to the cerebellum were obtained. As clusters relating to the superior cerebellum may be erroneously attributed to the ventral aspect of the fusiform cortex, a broad region of interest (ROI) surrounding the cerebellum was selected to identify possible relevant volumetric differences (MNI x/y/z: -60 to 60/-30 to -90/-5 to -60). Additionally, publications occasionally used vague terminology to report implicated regions of the cerebellum. To specify which regions were affected in both states of anorexia and obesity relative to healthy controls, the *spatially unbiased atlas template of the cerebellum and brainstem (SUIT)*[1] was used. FWE-corrected clusters produced by GingerALE were visualised on *MANGO*[2], with cluster-based ROI findings identified using the "nearest grey matter (MNI)" *MANGO* template, and compared with *SUIT*. Using an MNI template file, the output FWE-corrected clusters generated from GingerALE were inserted as a *MANGO* overlay to visualise regions of anatomical difference.

Prior to conjunction analyses, a logical overlay was inserted over datasets to identify any regions of overlap regarding decreased cerebellar volume. Analyses were performed by incorporating the AN/BN, OB and normative output result files (NIFTI format) as input files within the GingerALE GUI. Data was uncorrected, set to 1,000 permutations (as individual analyses were FWE corrected) and had a cluster significance threshold of $p<0.01$.

#### Table S1. Regional findings and coordinates from collected all studies

| First Author | Effect | Hemisphere | Coordinates (x, y, z) | Cluster Extent (mm$^3$/voxels.) | Z/T/p Value | Covariates (#; Covariate Type) |
|---|---|---|---|---|---|---|
| **[a] AN/BN (n=11 papers; 23 coordinates)<br>AN-only (n=8 papers; 16 coordinates)** | | | | | | |
| *Lenhart, 2022* | HC>AN | L | -33, -74, -48 | 2062 voxels | T=7.2 | 1; BMI |
| | | R | 36, -45, -45 | 2003 voxels | T=6.8 | |
| *Mishima, 2021* | HC>AN | R | 21, -69, -41 | N/A | T=5.77 | 3; AGE, TIV, Total GMV |
| *Phillipou, 2018* | HC>AN | R | 21, -63, -41 | 985 voxels | T=4.37 | 0 |
| [a] D'Agata, 2015 | HC>AN/BN | R | 24, -50, -26 | 85 voxels | T=2.95 | 5; AGE, BMI, etc. |
| | | L | -24, -54, -26 | 181 voxels | T=2.90 | |
| *Fonville, 2014* | HC>AN | R | 26, -56, -34 | 3229 | T=4.61 | 3; AGE, IQ, YEARS EDUCATION |
| | | L | -28, -56, -36 | 1573 | T=5.03 | |
| | | R | 28, -50, -16 | N/A | T=4.16 | |
| [a] *Amianto, 2013* | HC>AN/BN | L | -36, -48, -36 | 657 | p=0.001 | 3; AGE, TIV, DIAG. |
| | | R | 52, -70, -28 | 441 | p=0.001 | |
| | | L | -24, -22, -32 | 104 | p<0.001 | |
| | | R | 22, -62, -12 | 71 | p<0.005 | |
| *Bomba, 2013* | HC>AN | R | 23, -82, -45 | 10,447 | T=6.51 | 5; AGE, BMI, AGE AN ONSET, AN DURATION, PRIMARY/SECONDARY AMENORRHEA |
| *Brooks, 2011* | HC>AN | L | -4, -54, -26 | N/A | T=3.11 | 2; AGE, TOTAL GMV |
| *Boghi, 2011* | HC>AN | R | 35, -40, -54 | 5707 voxels | T=4.92 | 2; AGE, ICV |
| | | L | -17, 58, -35 | 299 voxels | T=3.84 | |
| | | R | 40, -61, -22 | 6409 voxels | T=6.11 | |
| | | L | -30, -55, -15 | 2315 voxels | T=5.28 | |
| *Gaudio, 2011* | HC>AN | L | -4, -81, -29 | 250 | T=4.63 | 1; AGE |
| | | L | -20, -47, -31 | 85 | T=3.53 | |
| | | L | -20, -68, -35 | 45 | T=3.34 | |
| [a] *Joos, 2010* | HC>AN/BN | R | 42, -73, -41 | 38 | T=4.34 | 0 |
| **OB (n=6 papers; 22 coordinates)** | | | | | | |
| *Shan, 2019* | HC>OB | R | 51, -57, -41 | 181 | T=7.37 | 3; |
| | | R | 51, -62, -29 | 322 | T=6.44 | |
| | | R | 38, -74, -12 | 81 | T=5.95 | |
| *Wang, 2017* | HC>OB | L | -27, -37, -47 | N/A | T=3.03 | 1; |
| | | L | -24, -73, -47 | N/A | T=2.94 | |
| *Ou, 2015* | HC>OB | L | -20, -78, -35 | N/A | T=5.22 | 2; AGE, SEX |
| *Jauch-Chara, 2015* | HC>OB | R | 15, -72, -50 | N/A | T=4.81 | 1; |
| | | L | -30, -60, -26 | N/A | T=5.23 | |
| | | L | -15, -67, -60 | N/A | T=4.25 | |
| | | R | 15, -70, -50 | N/A | T=5.31 | |
| | | R | 20, -52, 20 | N/A | T=N/A | |
| | | L | -32, -57, -27 | N/A | T=3.74 | |
| *Dommes, 2013* | HC>OB | R | 11, -39, -4 | 245 voxels | T=11.54 | 0 |
| | | L | -12, -35, -6 | 156 voxels | T=10.0 | |
| | | L | -27, -54, -34 | 36 voxels | T=6.60 | |

| | | | | | | |
|---|---|---|---|---|---|---|
| | | L | -18, -54, -28 | >5 voxels | T=5.31 | |
| | | R | 56, -63, -32 | N/A | T=6.01 | |
| | | R | 54, -60, -47 | N/A | T=5.99 | |
| *Mueller, 2012* | HC>OB | R | 36, -64, -33 | N/A | T=5.2 | 2; AGE, SEX |
| | | L | -31, -6, -30 | N/A | T=5.29 | |
| | | L | -52, -54, -38 | N/A | T=3.79 | |
| | | L | -45, -46, -44 | N/A | T=3.4 | |
| **NOR (n=9 papers, 10 studies; 49 coordinates)** | | | | | | |
| | | R | 44, -73, -29 | 4845 voxels | T=2.32 | |
| | | R | 26, -69, -29 | ^ | T=2.28 | |
| *Weise, 2019a* | -CORR (TWIN) | R | 32, -81, -26 | ^ | T=2.25 | 1; SEX |
| | | L | -30, -57, -45 | 799 voxels | T=2.28 | |
| | | L | -20, -61, -47 | ^ | T=1.96 | |
| | | L | -22, -72, -41 | ^ | T=1.74 | |
| | | R | 26, -81, -24 | 1537 voxels | T=2.07 | |
| | | R | 44, -72, -30 | ^ | T=2.01 | |
| | | R | 36, -76, -29 | ^ | T=2.01 | |
| *Weise, 2019b* | -CORR | R | 42, -51, -50 | 495 voxels | T=1.98 | 1; AGE |
| | | R | 36, -76, -29 | ^ | T=1.86 | |
| | | R | 27, -70, -45 | ^ | T=1.75 | |
| | | L | -34, -60, -47 | ^ | T=1.98 | |
| | *+CORR* | *R* | *11, -37, -47* | *NOT ASSESSED* | *Z=3.46* | |
| | | *R* | *33, -57, -41* | *NOT ASSESSED* | *Z=3.25* | |
| *Yao, 2016* | -CORR | L | -8, -79, -48 | 994 voxels | Z=4.24 | 4; AGE, GENDER, HANDEDNESS, GLOBAL GMV |
| | | R | 6, -79, -41 | 994 voxels | Z=3.97 | |
| | | L | -33, -57, -63 | 158 voxels | Z=3.52 | |
| | | R | 38, -65, -60 | 37 voxels | Z=3.26 | |
| *Figley, 2016* | -CORR | R | 8, -7, -5 | 5367 voxels | p<0.001 | 0 |
| | | R | 31, -68, -38 | 9939 | T=7.18 | |
| *Masouleh, 2016* | -CORR | L | -25, -73, -32 | 9984 | T=6.82 | 2; AGE, SEX |
| | | L | -30, -78, -17 | 5100 | T=6.1 | |
| | | R | 36, -72, -18 | 862 | T=6.40 | |
| | | R | 36, -54, -54 | 996 | T=4.46 (HT) | |
| | | R | 36, -54, -54 | 996 | T=4.46 (HT) | |
| | | R | 36, -54, -54 | 996 | T=4.46 (HT) | |
| | | R | 38, -33, -30 | 8 | T=4.46 (HT) | |
| | | R | 36, -54, -54 | 996 | T=4.46 (HT) | |
| | | R | 27, -70, -42 | 13 | T=4.46 (HT) | |
| *Janowitz, 2015* | -CORR | R | 6, -78, 28 | 6159 | T=4.46 (HT) | 5; AGE, BMI, AGE AN ONSET, AN DURATION, PRIMARY/SECONDARY AMENORRHEA |
| | | R | 6, -78, 28 | 6159 | T=4.46 (HT) | |
| | | R | 6, -78, 28 | 6159 | T=4.46 (HT) | |
| | | L | -33, -46, -50 | 24 | T=4.46 (HT) | |
| | | L | -23, -45, -51 | 1 | T=4.46 (HT) | |
| | | R | 6, -78, 28 | 6159 | T=4.46 (HT) | |
| | | R | 6, -78, 28 | 6159 | T=4.46 (HT) | |
| | | R | 24, -34, -11 | 138 | T=4.46 (HT) | |

| | | | | | | |
|---|---|---|---|---|---|---|
| | | R | 38, -33, -30 | 8 | T=4.46 (HT) | |
| | | L | -35, -55, -11 | 6 | T=4.46 (HT) | |
| | | L | -26, -28, -21 | 27 | T=4.46 (HT) | |
| | | R | 18, -66, -30 | N/A | p=0.0174 (FDR) | |
| *Kurth, 2013* | -CORR | L | -15, -72, -24 | N/A | p=0.0174 (FDR) | 2; AGE, SEX |
| | | R | 26, -65, -36 | N/A | p=0.0174 (FDR) | |
| | | L | -38, -33, -24 | N/A | p=0.0389 (FDR) | |
| *Weise, 2013* | -CORR | L | -8, -84, -38 | 405 voxels | p=0.013 | 3; AGE, SEX, HANDEDNESS |
| | | R | 50, -50, -31 | 108 | T=3.86 | |
| *Walther, 2010* | -CORR | L | -49, -47, -33 | 250 | T=3.31 | 1; HYPERTENSION |
| | | R | 16, -66, -32 | 12383 | T=5.13 | |

***Table S1.*** *Demographics of all studies used for analysis, including exclusion covariates, cluster extent measurements and MNI/Talairach coordinates; [a] – Publication used in AN/BN meta-analysis but excluded from exploratory AN-only analysis (n=8).*

*[**Abbreviations**: AN – Anorexia Nervosa; BMI – Body Mass Index; BN – Bulimia Nervosa; CORR – Correlation; DIAG. – Diagnosis; FDR – False Discovery Rate; GMV – Grey Matter Volume; HC – Healthy Control; HT – Height Threshold; ICV – Intracranial Volume; IQ – Intelligence Quotient; L – Left; N/A – Not Available; NOR – Normative; OB – Obesity; R – Right; TIV – Total Intracranial Volume]*

#### 3.0 – Results

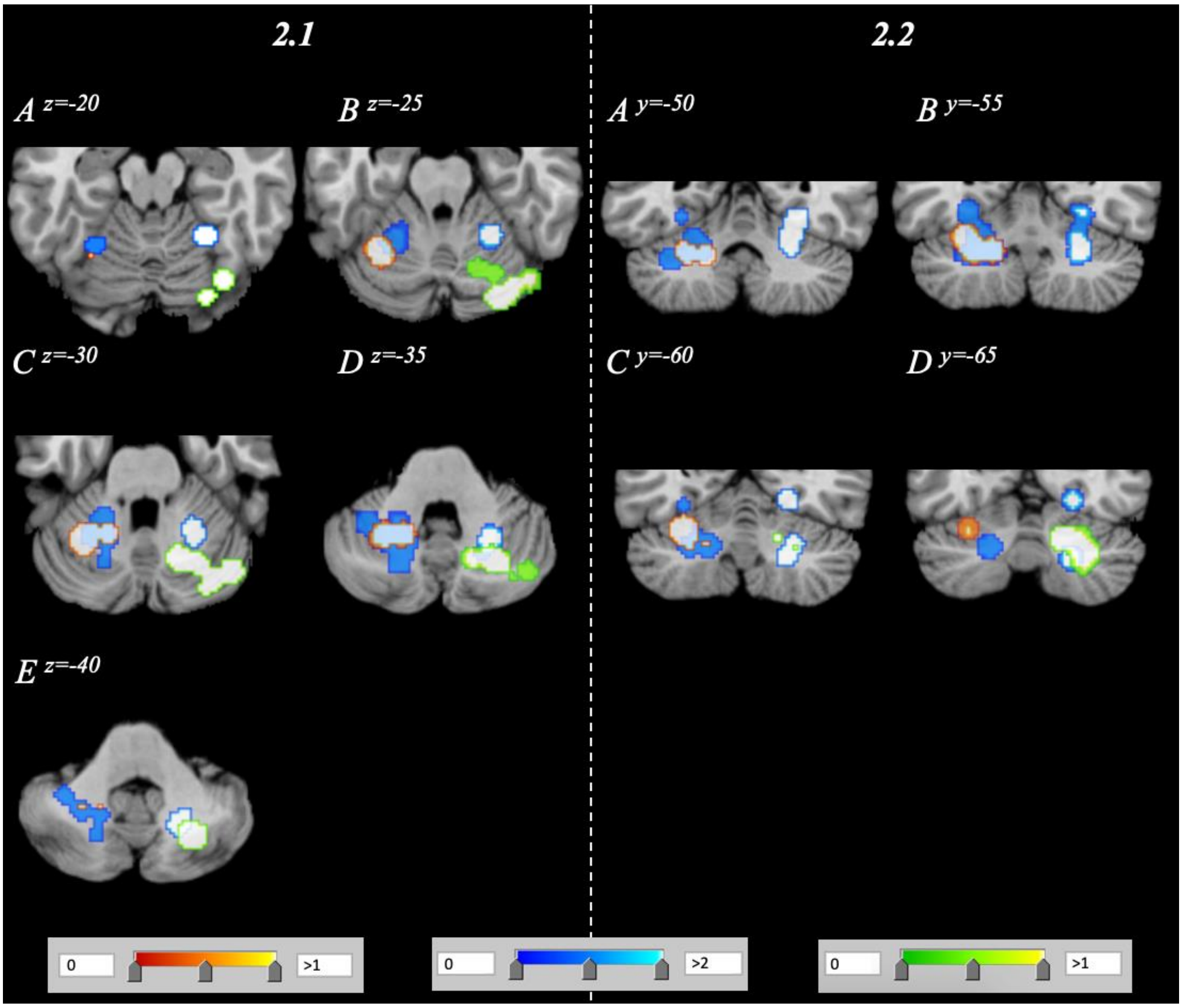


***Figure S2.*** *Pooled visualisation of volume reduction for the following three analyses; AN/BN vs. HC (blue); OB vs. HC (red/orange); Normative sample analysis (green). Image encompasses the entire cerebellum with a range of z=-20-z=-40 and y=50-y=-65 in axial (**S2.1**) and coronal (**S2.2**) orientations.*

**Table S2. Omission-by-one-publication jackknife analyses within OB and NOR findings**

| **OB – Omitted Publication** | **L CB A** | **L CB P** |
|---|---|---|
| *Shan, 2019* | Y | Y |
| *Wang, 2017* | Y | Y |
| *Ou, 2015* | Y | Y |
| *Jauch-Chara, 2015* | N | N |
| *Dommes, 2013* | N | N |
| *Mueller, 2012* | N | N |
| *OB Repeatability:* | *3/6* | *3/6* |
| **NOR – Omitted Publication** | **R CB P** | - |
| *Weise, 2019(a)* | Y | - |
| *Weise, 2019(b)* | Y | - |
| *Huang, 2019* | Y | - |
| *Yao, 2016* | Y | - |
| *Figley, 2016* | Y | - |
| *Masouleh, 2016* | Y | - |
| *Janowitz, 2015* | Y | - |
| *Kurth, 2013* | Y | - |
| *Weise, 2013* | Y | - |
| *Walther, 2010* | Y | - |
| *NOR Repeatability:* | *10/10* | - |
| **AN/BN – Omitted Publication** | **L CB P/A**[a] | **R CB A/P** [a] |
| *Lenhart, 2022* | Y | Y |
| *Mishima, 2021* | Y | Y |
| *Phillipou, 2018* | Y | Y |
| *D'Agata, 2015* | Y | N |
| *Fonville, 2014* | Y | N |
| *Amianto, 2013* | Y | Y |
| *Bomba, 2013* | Y | Y |
| *Brooks, 2011* | Y | Y |
| *Boghi, 2011* | Y | Y |
| *Gaudio, 2011* | Y | Y |
| *Joos, 2010* | Y | Y |
| *AN/BN Repeatability:* | *11/11* | *9/11* |
| **AN-only – Omitted Publication** | - | **Identified Post-Omission** |
| *Lenhart, 2022* | - | Y; L CB P |
| *Mishima, 2021* | - | N |
| *Phillipou, 2018* | - | Y; L CB P |
| *Fonville, 2014* | - | Y; L CB P/A[a] |
| *Bomba, 2013* | - | Y; L CB P |
| *Brooks, 2011* | - | N |
| *Boghi, 2011* | - | N |
| *Gaudio, 2011* | - | N |
| *AN Repeatability:* | - | *4/8* L CB P*; 1/8* L CB P/A[a] |

**Table S2.** Omission-by-one-publication jackknife analyses within OB and NOR, and AN/BN publications to assess whether cerebellar findings are (Y) or are not (N) present upon re-assessment. A sensitivity analysis was also conducted for the exploratory AN-only cohort dataset, despite no cluster-based findings identified.

[a] – Findings including relative even distributions (within a 10% difference) between anterior and posterior cerebellar volume are noted together, with the lobe containing higher finding-based percentage allocation reported first.

[**Abbreviations**: AN – Anorexia Nervosa; A – Anterior Lobe; BN – Bulimia Nervosa; CB – Cerebellum; L – Left; N – No; NOR – Normative; OB – Obesity; P – Posterior Lobe; R – Right; Y – Yes]
